# An Open Demonstrator for an Interoperable Clinical Decision Support System for the Detection of Systemic Inflammation and Sepsis in Pediatric Intensive Care

**DOI:** 10.64898/2026.08.04.26359683

**Authors:** Marcel Schack, Henning Rathert, Julia Böhnke, Nicole Rübsamen, Louisa Bode, André Karch, Mohammad Khair Almekkawi, Michael Marschollek, Philipp Beerbaum, Antje Wulff, Thomas Jack

**Affiliations:** Peter L. Reichertz Institute for Medical Informatics of TU Braunschweig and Hannover Medical School, Karl-Wiechert-Allee 3, 30625 Hannover, Germany; CAIMed – Lower Saxony Center for Artificial Intelligence and Causal Methods in Medicine, Appelstr. 9a, 30167 Hannover, Germany; Department of Pediatric Cardiology and Intensive Care Medicine, Hannover Medical School, Carl-Neuberg-Str. 1, 30625 Hannover, Germany; Institute of Epidemiology and Social Medicine, University of Münster, Germany, Albert-Schweitzer-Campus 1, 48149 Münster, Germany; Big Data in Medicine, Department of Health Services Research, School of Medicine and Health Sciences, Carl von Ossietzky Universität Oldenburg, Oldenburg, Germany

**Keywords:** pediatric intensive care, CDSS, sepsis, SIRS, open demonstrator, dataset, DTA

## Abstract

**Background:** Sepsis is a life-threatening condition triggered by infection and associated with dysregulated immune response of the patient followed often by multiorgan dysfunction or failure. In the clinical evolution of sepsis towards organ dysfunction, early initiation of a suited therapy significantly increases patient outcomes and reduces mortality rates. Since electronic health records provide data in a machine-readable format, this process could be supported by computerized systems.

**Methods:** We developed an interoperable, time-sensitive CDSS that able to detect systemic inflammation and the different classifications of sepsis (bacterial/viral, suspected/proven, on admission/PICU acquired) in pediatric patients based on the analysis of routine clinical data. This application is provided as part of this publication as an open demonstrator (web application), and the usability and accuracy of the CDSS is shown by a retrospective creation of sepsis outcome labels for a routine data set of 4,655 pediatric patients. As a reference standard, the patients were manually assessed by blinded clinical experts.

**Results:** In comparison with the reference standard, the CDSS achieved sensitivity of 96.9% (95% CI: 80.9-99.6%) and specificity of 99.1% (95% CI: 95.1-99.8%). In the context of a sepsis outcome labeling for 4,655 patients, the CDSS detected 4,342 episodes of inflammation of which 1,723 were classified as sepsis.

**Conclusions:** We demonstrated that our routine-data based CDSS is able to perform a complex sepsis detection process with high diagnostic accuracy. Such CDSS with the ability to differentiate between SIRS, sepsis on admission, suspected and proven sepsis can prospectively support clinical management, monitoring and quality management.

## 1. BACKGROUND

Sepsis is a life-threatening condition triggered by a dysregulated immune reaction to harmful microorganisms [1]. In 2017, there were 48.9 million sepsis cases worldwide, particularly 20.3 million incidents and 2.9 million deaths among pediatric patients younger than five years [2]. In the clinical evolution of sepsis towards organ dysfunctions or failure, early initiation of an effective therapy significantly increases patient outcomes and reduces mortality rates [3,4]. In the case of a particularly severe infection, the immune response can affect the entire body system, leading to an exaggerated immune reaction that potentially affects all organ systems. Diagnostic and medical decision-making is a challenging and recurring task for clinicians, especially in areas with elevated sepsis risk in combination with a highly dynamic area of work such as in pediatric intensive care [5,6]. To recognize sepsis in children, age-specific diagnostic criteria have been developed that take into account the patient’s vital signs and laboratory values. In the evolution of the definition of pediatric sepsis from the IPSCC criteria [7] to the Phoenix criteria [8], the focus has steadily shifted from inflammation to sepsis-associated organ dysfunction, mainly to improve the specificity and clinical relevance of the results of this diagnostic approach. Furthermore, this shift fails to take into account that the onset of sepsis is marked in its earliest stage by the onset of inflammation. This inflammatory response is defined by an increase in white blood cell count, fever (mandatory criteria), tachypnea, and tachycardia (optional criteria), and are the diagnostic focus of the 2005 IPSCC criteria [7]. Therapeutic measures initiated as early as this (pre-)phase of sepsis can prevent the subsequent development of organ dysfunction or failure and be of the greatest benefit to the critically ill child. A recent publication addressing the development of a sepsis prediction model using the newly defined Phoenix criteria clearly demonstrated that the focus on the presence of organ dysfunction in the Phoenix sepsis criteria should be critically questioned. The authors once again clearly emphasize that the Phoenix score was not developed for early screening for sepsis and treatment prior to the onset of organ dysfunction. This is also reflected in the results of this cohort, which show a comparatively high mortality rate in the group of patients with sepsis defined based on the Phoenix criteria [9]. This should be taken into account, particularly in the development of clinical decision support systems (CDSS) and predictive models based on machine learning, which aim to predict diagnoses as early as possible and before the onset of clinical symptoms. This should be taken into account in particular when developing clinical decision support systems (CDSS) and prediction models based on machine learning, which aim to predict diagnoses as early as possible and before the onset of the clinical symptoms.

The quality of clinical decision-making depends on the multimodal information available on the patient’s state, but the collectability, quality and speed of data representation have also major significance. Unfiltered information, the time pressure of the clinical routine and the complexity of the different pediatric diseases all contribute to clinical decisions with a lower quality than desired [5]. To overcome this challenge, clinicians can be supported by CDSSs. By reusing and valorizing clinical routine data, CDSS can provide relevant information, guide clinicians in their decision-making, and detect diseases. However, multiple use of clinical routine data is often complicated due to vendor-dependent, proprietary, and heterogeneous data sources, with an inherent lack of standardization or interfaces. For the development of a CDSS for clinical routine that is not designed as a stand-alone system, it is necessary to harmonize the underlying routine data and transfer them into interoperable formats [10,11]. In the context of medical data, vendor-neutral interoperable standards such as HL7 CDA/CCR, HL7 FHIR, OMOP, or openEHR are used along with ontologies or terminologies [12]. Since the integration of a CDSS component into existing systems requires hospital-specific adjustments, the use of interoperable standards can speed up this cumbersome process. Intensive care medicine, with its need for many time-critical decisions, requires applications that can be fully integrated into the respective processes without major additional effort and, if possible, based on routine data. Although the use of a CDSS can improve patient outcome, recent research on sepsis detection with interoperable CDSS in PICU is scarce [13]. Related approaches published by Alturki et al. [14], Dewan et al. [15], Scott et al. [4], Vidirne et al. [16], Eisenberg et al. [17], Le et al. [18], Sepanski et al. [19] and Cruz et al. [20], are not focusing on interoperable standards. Furthermore, existing approaches are not publicly available for other researchers.

In our previous work, we designed knowledge-based CDSSs for detection of pediatric SIRS, hematologic organ dysfunction or acute kidney injury that uses routine data transformed in interoperable formats using openEHR [21–24]. Furthermore, we proposed a data integration pipeline that resulted in an interoperable dataset with over 4,000 pediatric patients [25].

In this work, we strive to further develop these knowledge-based CDSSs for the detection of sepsis in pediatric patients. We first adapt the already developed CDSS for the detection of a SIRS by extending the underlying rule system for sepsis criteria and evaluating its diagnostic test accuracy (DTA) for longitudinal patient data [26]. In a second step, we develop an interoperable CDSS capable of detecting sepsis in pediatric patients from routine data. This procedure is derived from the medical background where SIRS serves as a foundation for sepsis detection. Next, we present the release of our CDSS as an open demonstrator.

Finally, we use the CDSS to retrospectively generate outcome labels for an available unique dataset from our setting (see above). We hypothesize that this step will enable the use of machine learning approaches for our labeled dataset and for comparable datasets from other institutions in the near future. By publishing our CDSS as an open demonstrator, we strive to enable other researchers to use the system and to improve the performance of machine learning algorithms for sepsis research purposes.

## 2. METHODS

For the implementation of our rule-based CDSS for the detection of sepsis, we fully rely on the successfully evaluated architecture of our published CDSS for SIRS detection [21]. We stick to the key characteristics and published architecture of our CDSS. For details, we refer to Wulff et al. [21] and Bode et al. [23]. Before utilization, we identified potentials of improving this SIRS-CDSS by refining the developed rule base and expanding the used database. Previously, we used clinical routine best practices and agreed-upon clinical diagnostic guidelines as a basis for our SIRS rule base. However, as reported by Wulff et al. [21], the implementation of specific aspects was limited by missing or inaccessible data. Through our efforts in integrating an interoperable data set [25], the implementation of more advanced rules is now feasible. In this context, we reviewed the implemented rules with experienced clinical experts and improved these with more fine-grained adaptations. Four our new use case of sepsis detection, we implemented an iterative step-by-step approach.

### Step 1: Data Identification and Hosting

The first step involves the identification of data assets relevant for the detection of pediatric sepsis from various data sources. Most data are routinely stored in the local patient data management system *m.life*^2^ by company medisite of the PICU at Hannover Medical School (MHH). Additional heterogeneous and proprietary data sources from individual, isolated data silos within the hospital have also been incorporated. After an interdisciplinary assessment of the necessary data with two experienced intensive care pediatricians, we developed data integration processes. Data was stored in an interim SQL-database before being transferred with the open-source tool HaMSTR [27] an openEHR-based clinical data repository called *ehrbase*^3^. For further details, we refer to Bode et al. [25].

### Step 2: Knowledge Acquisition and Modeling

For the representation of knowledge for this new use case, we adopted our previous approach, which involves the use of commonKADS as a methodology for acquiring, modelling, and processing knowledge [28]. We performed interviews with two experienced intensive care pediatricians for knowledge acquisition and transfer. We then developed human-readable rules for diagnosing sepsis in the PICU were developed, that were transformed into machine-readable code by computer scientists (see step 4). In preparation for the evaluation, the clinicians created a reference standard for the occurrence of sepsis for the 168 patients in our study cohort [29].

### Step 3: Rule Development

We used the Business Rules Management System Drools by JBoss (Red Hat) [30] to transfer expert knowledge into computable code that we run on the prospective, monocentric, double-blinded, diagnostic CADDIE2 study cohort (n=168 pediatric patients) [29]. Independently, two experienced pediatricians created a reference standard for evaluating the sepsis CDSS by manual chart review of patient’s medical records, considering the SIRS and sepsis diagnostic criteria. If, after this review, pediatricians came up with different results, these cases were resolved through detailed discussion until a consensus was reached. For the development process, pediatricians and computer scientists optimized the rule base or the reference standard by reviewing and discussing the causes of false positive and false negative alarms.

### Step 4: Demonstrator Implementation

To provide our implemented CDSS to other researchers and show its functionality, we developed a publicly accessible open demonstrator. The open demonstrator mimics the CDSS’s reasoning process and displays its functionalities. To comply with local data security regulations and protect patient information, we manually created synthetic data as a basis for our open demonstrator. The aim of the manual synthetization is to provide use cases for all outcome labels and patient histories.

### Step 5: Generation of Clinical Outcome Label

Finally, we used our newly developed CDSS to retrospectively create outcome labels for sepsis to a cohort of 4,655 patients, whose medical data were not involved in the development process. In addition, we prepared this dataset to make it available to other researchers, promoting open science and addressing the scarcity of publicly available annotated datasets for critically ill children [31].

### Statistical Analysis and Diagnostic Test Accuracy Estimation

For each index test (i.e., the detection models; here the SIRS CDSS and the sepsis CDSS), we merged patient data (age, sex, and observation periods, i.e. length of stay at PICU) with start and end times of diagnostic episodes according to the reference standard (clinician’s diagnoses blinded to index test results) and respective index test. Per patient, any episodes less than 36 hours apart were merged into one continuing episode. There were neither missing nor indetermined index-test results.

We estimated DTA metrics—sensitivity, specificity, and their 95% Wald confidence intervals with the method by Böhnke et al. [26]: We labelled (i.e., true positive, false positive, false negative, or true negative) the dataset using hour as time unit to estimate the DTA at both the time-level (measuring the model’s precision in identifying correct time points) and block-level (evaluating performance over clinically relevant time periods). Whenever there were no false positive or false negative results so that we could not estimate a 95% Wald confidence interval, we used Wilson’s method [32] to estimate a lower bound of DTA ignoring the data clustering (i.e. which DTA could have been achieved if even just one false result had occurred). We conducted exploratory stratified analyses for sex, age groups, and all combinations of sex by age groups, using the same methods as the main analyses (Appendix A). All analyses were performed in R version 4.4.2 (2024-10-31) [33]. To address the identified false-positive and false-negative episode labels, we carefully reviewed the erroneous episode labels to rule out any errors in knowledge acquisition, rule development, or methodological approach. We examined whether the entire sepsis episode identified by the CDSS was incorrect or whether the timing of the episode was incorrect when compared to the reference standard.

## 3. RESULTS

### Implementation of Sepsis Detection

Since a time-aware SIRS detection with a high accuracy is necessary to successfully detect sepsis, we developed the CDSS for the detection of sepsis as an extension of the SIRS-CDSS on top of its previously developed infrastructure. Our adapted rules for SIRS detection are described in Appendix B.

Analogous to our previous work, the internationally agreed-upon diagnostic criteria [7] and best practices from clinical routine serve as a foundation of the implemented rules. Figure 1 displays the reasoning process for the detection of sepsis including all adaptations. The left entry point indicates that only the presence of SIRS can start the reasoning process for the detection of sepsis. The structure of sepsis detection is a three-phase sequence consisting of a total of six rules, summarized in Table 1. In the first phase, the presence of proven sepsis is addressed by checking microbiological results from blood culture sampling for either one positive pathogen (P1a) or for two positive findings of common skin germs (P1b) within 72 hours to a SIRS episode. If either condition is true, the underlying SIRS episode is considered as a proven sepsis, and sepsis detection is ended. If neither of the two conditions are true, the underlying SIRS episode is classified as proven sepsis and sepsis detection ends.

**Figure 1.**
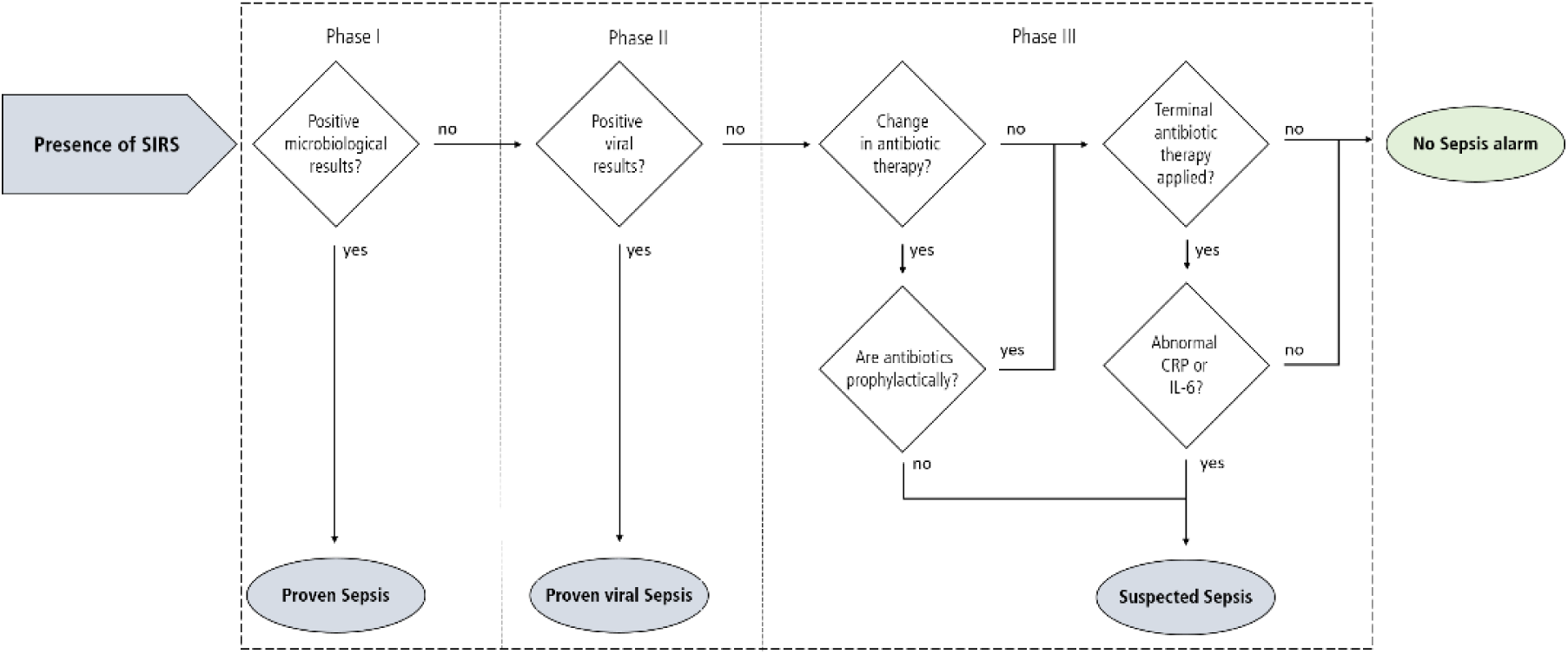
Simplified illustration of the three-phase reasoning process of the developed clinical decision support system for the detection of sepsis (not including details on timings). **Abbreviations:** SIRS, Systemic inflammatory response syndrome.

**Table 1.**
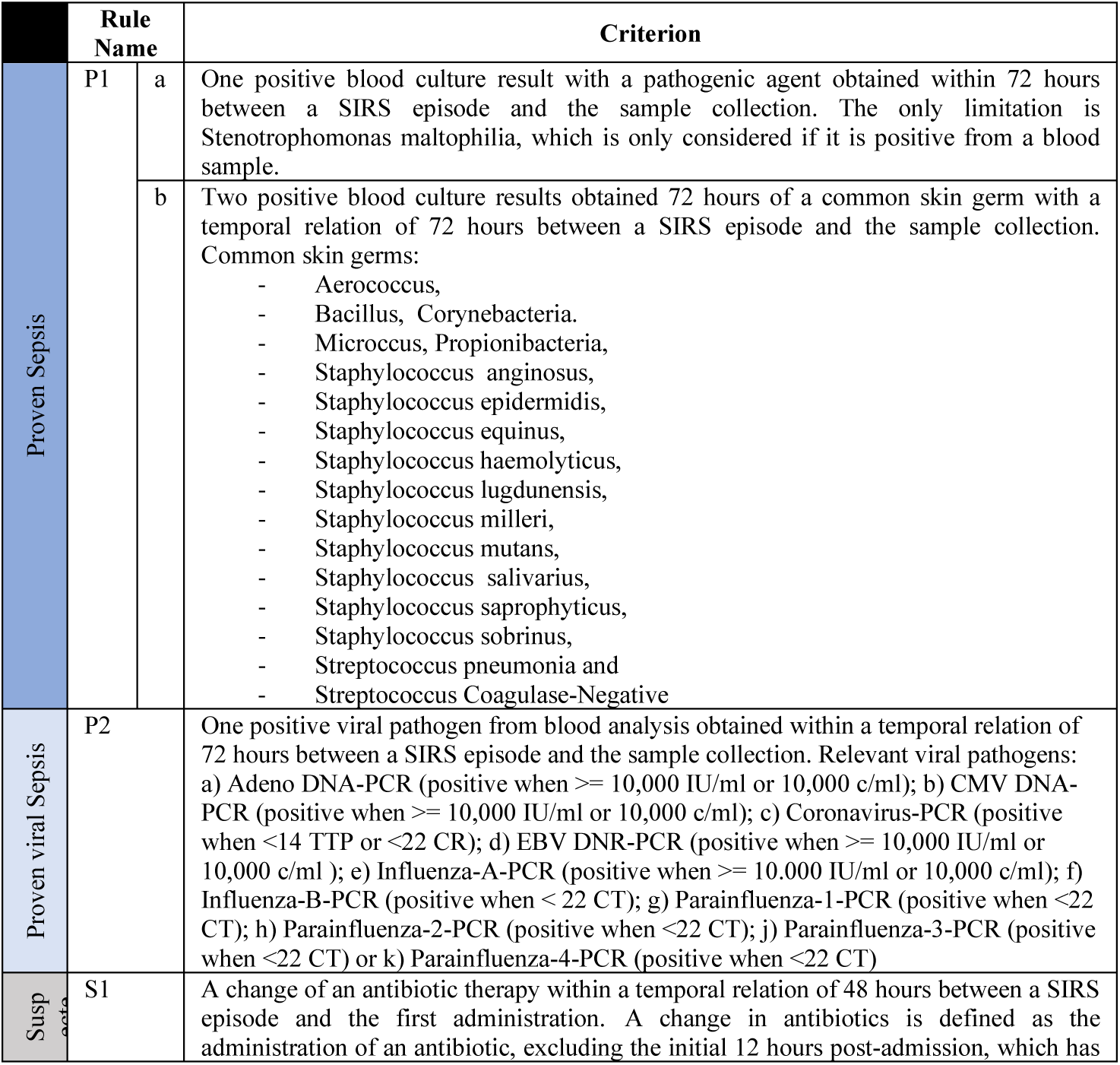
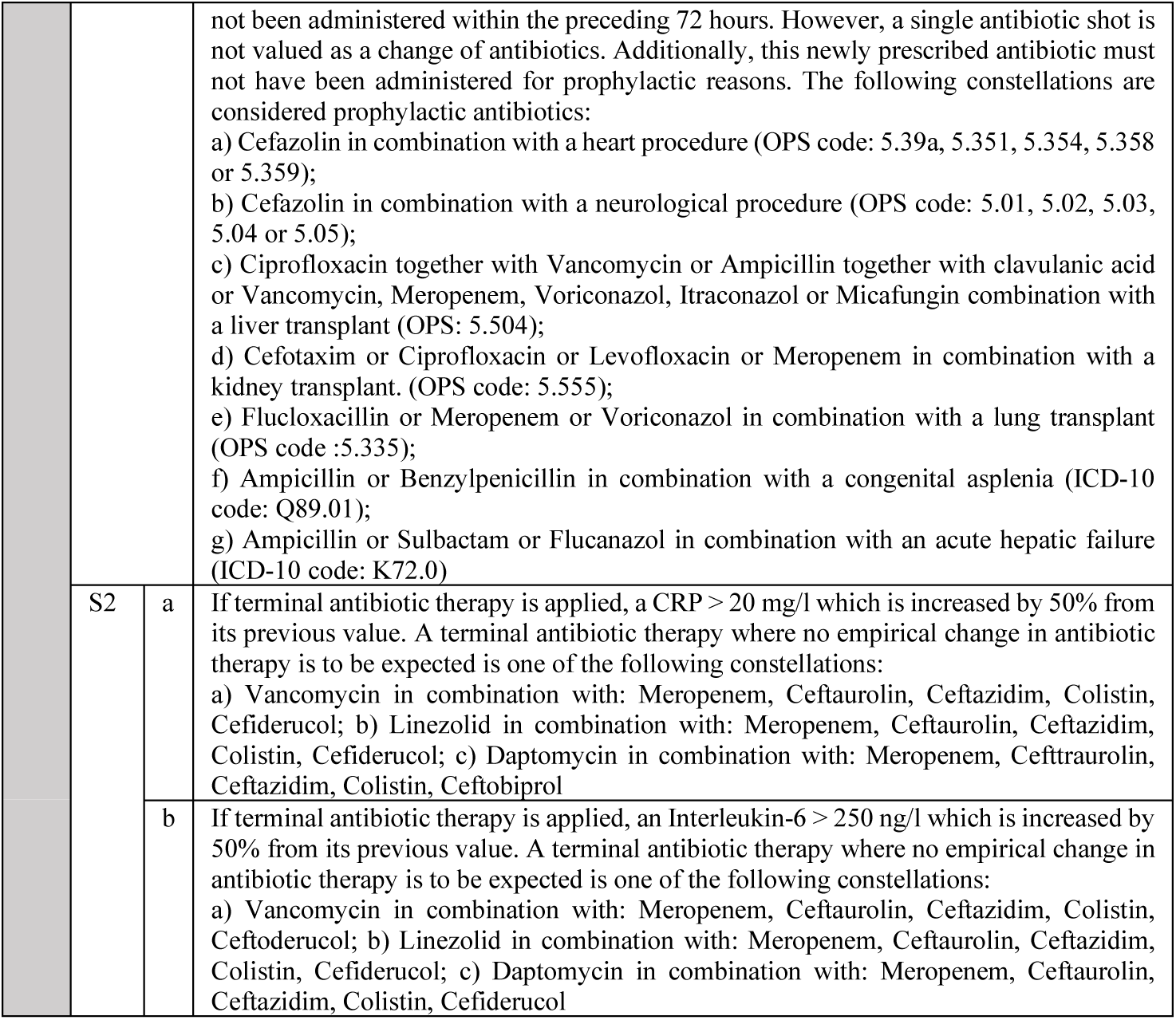
A summary of the rules that are currently in place within the clinical decision support system, which relate to the diagnosis of proven, proven viral and suspected sepsis. **Abbreviations:** CMV, Cytomegalovirus; CRP, C-reactive protein; DNA, Deoxyribonucleic acid; EBV, Epstein–Barr virus; ICD, International Classification of Diseases; OPS, Operationen-und Prozedurenschlüssel (Operation and Procedure Classification System), PCR, polymerase chain reaction; SIRS, Systemic inflammatory response syndrome.

If neither of the two conditions are satisfied, phase II checks for proven viral sepsis. A SIRS episode is labeled as a proven viral sepsis (P2) if a viral pathogen is detected in the laboratory results within 72 hours to a SIRS episode. The reasoning process concludes if the condition is true.

In the absence of proven viral sepsis detected by the CDSS, the third and final phase is triggered checking for suspected sepsis. Any change in antibiotic treatment within 48 hours in temporal correlation of a SIRS episode, which is not prophylactic due to a specific procedure or diagnosis, causes a suspected sepsis episode (S1). This rule intends to mimic how a pediatrician approaches an unconfirmed infection in clinical practice, which is addressed by newly prescribing or a change of antibiotic therapy. However, if the patient is given maximum escalated antibiotic therapy, a suspected infection will not be treated by changing the antibiotic therapy. In this case, the inflammation parameters CRP and interleukin-6 are analyzed. If this terminal antibiotic therapy is applied and either CRP is above 20 mg/l and has increased by 50% from its previous value (S2a) or Interleukin-6 is above 250 ng/l and has increased by 50% from its previous value (S2b), the underlying SIRS episode is also labeled as suspected sepsis. If none of the conditions of the three phases apply, the underlying SIRS episode does not become septic.

Provided that the outcome of a given test in one phase is either still pending or has not yet been conducted, the CDSS will proceed with the subsequent phases to facilitate a timely decision. The reasoning process is triggered by new findings or data on the patient, such as the result of a microbiological test. Consequently, this can result in an episode being elevated in severity from suspected to proven sepsis.

### Implementation of an Open Demonstrator

For providing an open demonstrator, we developed a web application hosted via GWDG Cloud Service from the University of Göttingen and the Max Planck Society^4^. The open demonstrator is available at: http://sepsis-cdss.plri.de.

To mimic the CDSS’s functionalities, we implemented a database hosting patient’s data. The focus of the demonstrator was not on interactive visualization design. We mimic the entire rule-based reasoning process and presents its results in a graphical user interface (GUI) and an explanation component, that guides users through the reasoning process. Upon launching the system, users must select a patient and a respective time point. Figure 2 displays the GUI implemented for that process within the open demonstrator. After patient selection, the implemented reasoning process is triggered and results are displayed automatically, such as in Figure 3, where the heart rate measured over time is represented by a red line. For comparison, the upper limit of the normal values for the respective age category of the patient is indicated as a black line. If the patient was equipped with a pacemaker, this would be indicated by a blue line. The phases in which the CDSS identifies the presence of sepsis or SIRS are indicated by an orange background.

**Figure 2.**
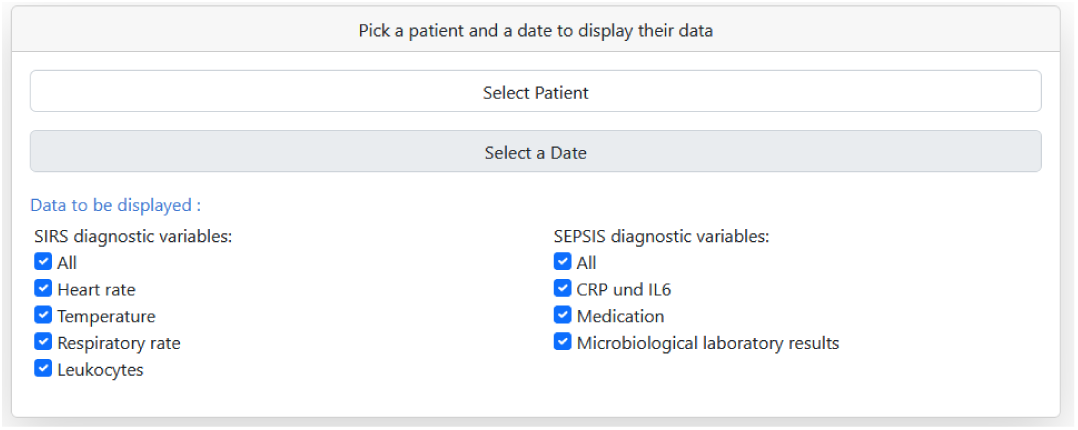
The graphical user interface allows the user to select the patient and the respective day for which the open demonstrator for the clinical decision support system is to be initiated. The demonstrator visualizes the functionality of the CDSS and is not intended to be used for medical purposes. Parameters can be explicitly selected or deselected using the provided checkboxes. **Abbreviations:** CRP, C-reactive protein; IL6, Interleukin 6, SIRS, Systemic inflammatory response syndrome.

**Figure 3.**
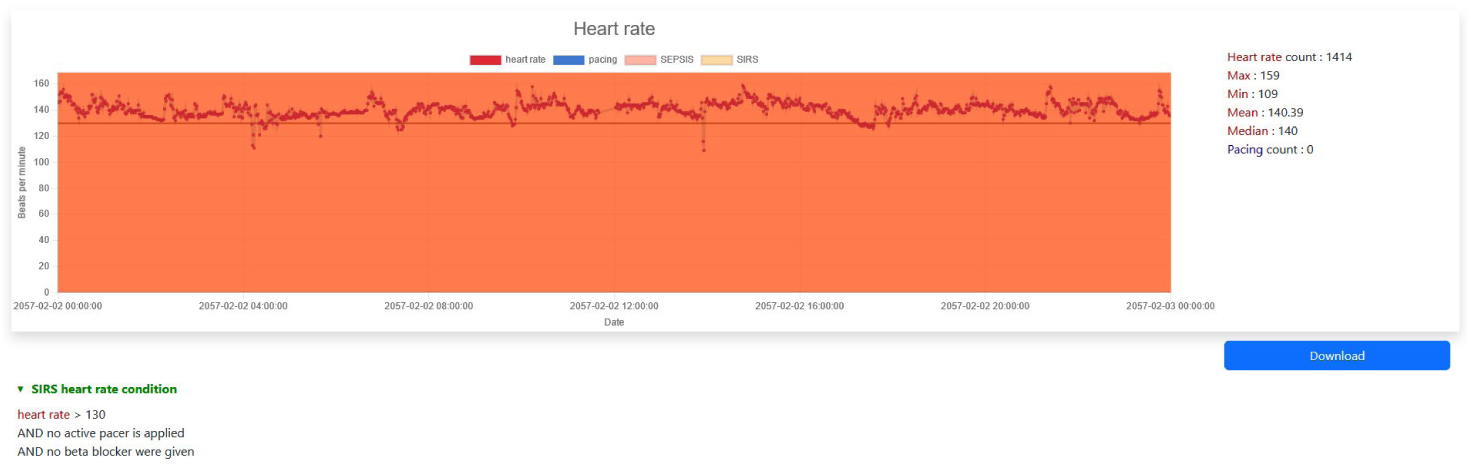
Exemplary output generated by the demonstrator for the purpose of displaying a heart rate. **Abbreviations:** SIRS, Systemic inflammatory response syndrome.

Minimum and maximum values, as well as median heart rate, are presented in a tabular format to the right of the diagram. The diagram is accompanied by a logical statement written in pseudo-code, which elucidates how the CDSS incorporates the specified parameter into its decision-making process.

Besides demographic information, up to seven separate diagrams display those decision-relevant vital signs and administered medications, to provide further understanding and explanation of the reasoning process and the outcome label. To save the provided results, the open demonstrator provides a export function of diagrams. Beneath every diagram and information, the explanation component provides whether the parameter is relevant to the outcome label.

#### Generation of clinical outcome labels

Finally, we used our developed CDSS to create retrospective outcome labels for SIRS and sepsis in an existing data set [31].

An overview of the labelling results is provided in Figure 4. In our dataset of 6,378 cases, from 4,655 different patients, the CDSS detected 4,342 SIRS episodes with a median age of 774 days on SIRS episode start (ranging from 0 to 6,558 days). Of these SIRS episodes, 1,723 episodes were classified as sepsis with a median age of 806 days, (range: 0 to 6,558 days). Specifically, there were 442 proven sepsis (median age at episode start: 790 days, range: 9 to 6,543 days), including 50 proven viral sepsis (median age at episode start: 1,453 days, range: 108 to 6,336 days), and 1,281 suspected sepsis episodes (median age at episode start: 828 days, range: 0 to 6,558 days).

**Figure 4.**
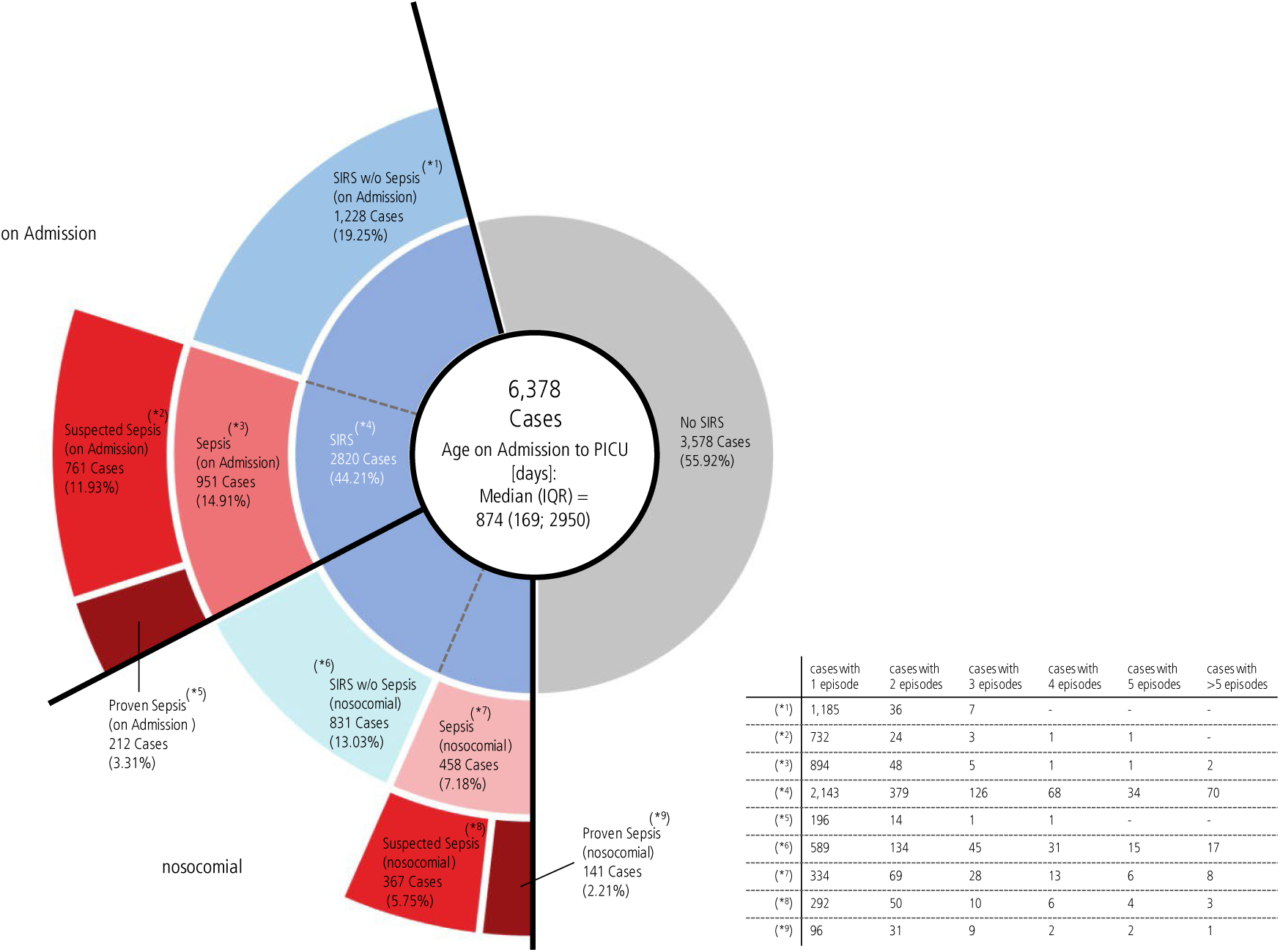
Overview of the label results. **Abbreviations:** Ep, Episodes; IQR, Interquartile range. SIRS, Systemic inflammatory response syndrome; Pat, Patients.

Regarding sex distribution, the sub-cohort suffering from SIRS consists of 1,044 females (45.19%) and 1,266 males (54.81%), and the sub-cohort suffering from sepsis had a relatively equal sex balance, with 509 females (45.81%) and 602 males (54.19%).

The cases illustrated in Figure 4 pertain to hospitalization. During a hospital stay, a patient may be admitted to the PICU several times and thus might have several cases of sepsis on admission.

Furthermore, it is important to note that a patient may develop both sepsis on admission and nosocomial sepsis at a later stage in their treatment.

### Evaluation and Statistical Analysis

The CDSS detected the presence of sepsis with sensitivities ≥96% and the absence of sepsis with specificities of ≥98% (Table 2) across the estimation levels. The model performed less well in certain clusters (i.e., newborns, neonates, and adolescents regardless of sex; see Appendix C).

**Table 2:**
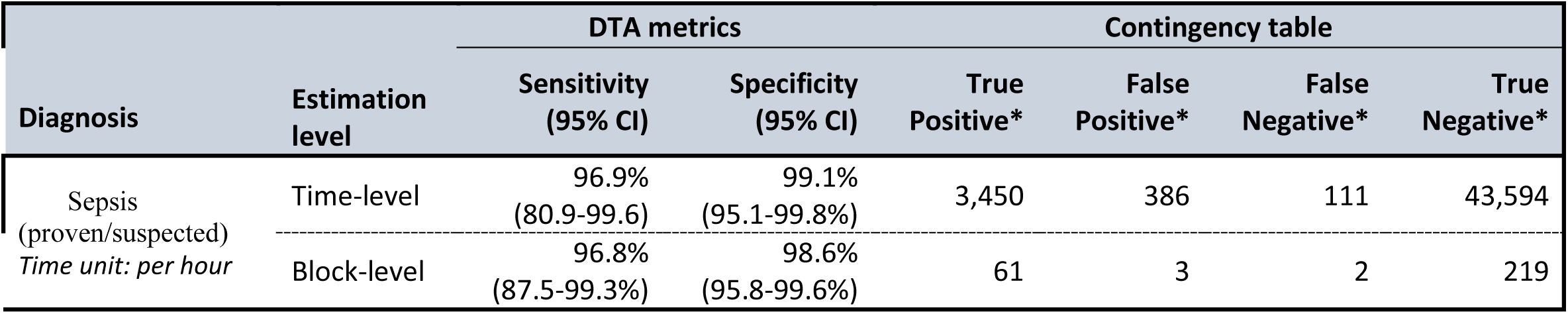
This table presents the DTA of the sepsis detection model (index test) as compared to the clinicians diagnostic decision, which defines the ground truth. **Abbreviations:** DTA, diagnostic test accuracy; CI; confidence interval

### Error Assessment

False positives (FP) or false negatives (FN) results occurred in only six out of 168 cases. All identified errors pertain to the complete absence or misrecognition of sepsis episodes. Each of the six erroneous cases involved a suspected sepsis episode and were attributable to rule S1 (Table 1). Erroneous cases No.1 and No.6 (both FN) were due to discordance between the interpretation of the clinical investigators, who defined the SIRS episodes in both cases as suspected sepsis. This conclusion was based on very subtle changes in the clinical state of the patient in combination with little changes of inflammation parameters. Since the extent of the increase in inflammatory parameters in both patients was not sufficient to trigger the rule, and as there were no changes following antibiotic therapy, both episodes were interpreted differently by the system and thus not recognized.

The second erroneous case (No.2) was caused by the CDSS incorrectly evaluating a change of antibiotic treatment. Specifically, ampicillin and sulbactam were administered for prophylactic treatment in relation to a superinfection, which is not the standard practice in the PICU. Such a combination of factors is not addressed by the CDSS’s established protocols.

The erroneous case No.3 was due to antibiotic therapy being escalated on a clinical basis in accordance with the liver-transplantation regimen. This would only be considered prophylaxis if the antibiotic had already been administered in the fixed combination for transplantations. Therefore, administered antibiotics can be defined as an escalation on a clinical basis in cases of suspected infection or sepsis.

Erroneous case No.4 resulted from a de-escalated antibiotics strategy due to decreasing inflammation parameters. A change in antibiotics is always regarded as fulfilled criterion (criterion S1) for a suspected sepsis by the CDSS. However, a scenario where the antibiotic strategy has been discontinued is not implemented.

In erroneous case No. 5, the administration of vancomycin was considered a change in antibiotic treatment, resulting in a false positive suspected sepsis episode. Vancomycin was added with a certain delay after admission of the patient to the PICU but was already initiated on the peripheral ward to treat a clinically new infection/sepsis. The CDSS is currently unable to adequately identify patients with existing sepsis on peripheral wards due to the unavailability of patient data management systems and digital EHR recording on these wards.

Two of the six errors identified were false negatives, resulting in a cumulative error period of 15 hours (17.6%) out of a total of 85 hours cumulative over all errors. The aggregate length of stay in the intensive care unit of all 168 patients utilized for the evaluation is 48,836 hours. Consequently, the CDSS erroneously categorizes 0.17% of the total period in question (Figure 5).

**Figure 5.**
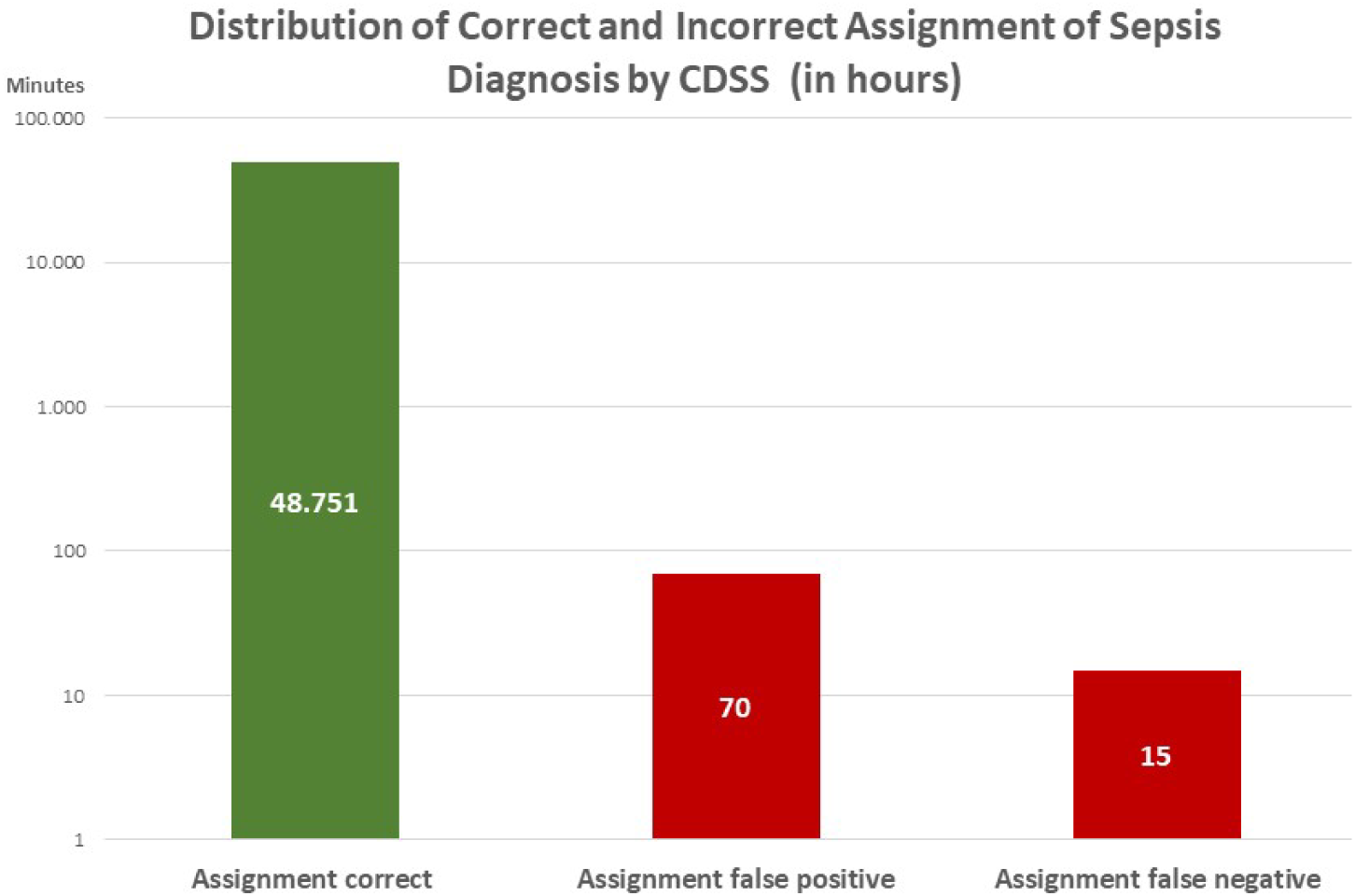
Distribution of correct and incorrect assignment of sepsis diagnosis by CDSS (in hours). **Abbreviations:** CDSS, Clinical Decision Support System.

## 4. DISCUSSION

Our approach offers a CDSS for the detection of inflammation and sepsis in critically ill pediatric patients and a publicly accessible open demonstrator. Our CDSS detected sepsis using well-established criteria by the International Pediatric Sepsis Consensus Conference (IPSCC) [7], achieving high sensitivities and specificities to detect the hour-based onset and the end of sepsis. Although alternative diagnostic criteria for the recognition of SIRS and sepsis exist in scientific literature [8], these have yet to be adopted in clinical practice. Accordingly, our system has been implemented with the IPSCC’s widely adopted clinical diagnostic criteria. The most important argument in favor of using the Goldstein IPSCC-sepsis definition for the diagnosis is above all to enable the system to detect an event as early as possible, which from a clinical point of view is the beginning of the systemic inflammation reaction and judge whether this reflects SIRS or sepsis. The most recent Phoenix criteria [8], based on organ dysfunction scoring, are not suitable to detect the clinical onset of the disease. Considering today’s times in which the machine learning-based development of sepsis prediction also seems feasible in children, the use of a detection model that can label the earliest possible time of clinical onset of sepsis, as reflected best by IPSCC criteria, is highly beneficial.

As already shown in our scoping review form the year 2019 [13], available data and research focusing on automated detection of pediatric SIRS and sepsis is still scarce and our presented model, based on real, routine clinical data for the detection of pediatric SIRS and sepsis, still represents innovative approach. Our findings showed that available data and research focusing on automated detection of pediatric sepsis was scarce. To the best of our knowledge, there is no other design that can be considered as equally reliable in terms of its accuracy, time sensitivity of labels and applicability.

The CDSS was evaluated in a sufficiently large pediatric patient cohort, using real clinical routine data from the PICU of MHH. Furthermore, to our knowledge, no other sepsis detection model for outcome labelling has been evaluated and provided as an open demonstrator. In alignment with our long-term objective of publishing an evolutionary longitudinal pediatric intensive care dataset [31], we will publish the retrospective outcome labels for SIRS and sepsis generated by our developed CDSS.

### Strengths and Limitation

On time level, our approach yielded a sensitivity of 96.9% and a specificity of 99.1% for sepsis. To further enhance our CDSS, we conducted a comprehensive analysis of the incorrectly classified episodes and formulated recommendations for future development. The recognition of alterations in antibiotic therapy in conjunction with patient transfers from other wards or hospitals represents a significant challenge for the CDSS, given the absence of pertinent data from external sources, particularly those outside the PICU. It would therefore be highly advantageous, in terms of infrastructure, to transfer patient data from other hospitals for clinical care and utilization in CDSSs.

A further challenge presented by the CDSS is the highly diverse patient cohort in the field of pediatric intensive care medicine, which is characterized by a broad range of underlying conditions. To accurately assess the efficacy of a change in antibiotic therapy in this complex domain, it is imperative that there is a heightened level of intensive collaboration between clinical domain experts and CDSS developers.

The data employed in the construction of the CDSS for sepsis detection and its conversion into an interoperable format were drawn from real-world, routine sources and had not been collected for this specific purpose. In a retrospective dataset, data quality is contingent upon the accuracy of the measurements and the entry of the data into the system. When introducing the system at other locations, it is of the utmost importance to make the requisite adjustments to the local systems, even if this process is facilitated by the choice of an interoperable format. This process of local implementation also requires clinical expertise to adapt rules that integrate, for example, the evaluation of treatment algorithms (antibiotics) or local specifics in patient monitoring. Nevertheless, our methodology offers a realistic estimation of the maximum DTA, thereby supporting the generalizability of our CDSS for sepsis in a real-world pediatric context. The exceptional DTA of our approach, coupled with the minimal incidence of false-positive alerts, renders it an optimal candidate for implementation, one that is readily accepted by users.

It is our intention to facilitate progress within the open science movement by providing an open demonstrator. The dataset and the outcome labels provided are ready for use with machine learning algorithms, allowing other researchers to adopt this approach directly.

### Future Directions

The next desired stage of the project would be, together with a certified manufacturer, the implementation of our approach as a real-time CDSS on PICU wards. We anticipate that the CDSS would have a significant impact on clinical practice and the management of sepsis in critically ill children by providing guidance to clinicians in their decision-making processes. Furthermore, we utilized our CDSS as a retrospective labelling system, enabling to retrieve a labelled dataset in a fraction of the time it would take a human, while maintaining comparable accuracy. The objective of retrospectively labeling a larger cohort is to provide a suitable dataset for machine learning approaches.

## Conclusion

We presented an automated CDSS capable of accurately detecting SIRS and sepsis using real-world data published as an open demonstrator. Moreover, we demonstrated that our approach successfully can be used to retrospectively generate precise outcome labels for existing real-world datasets in a time-efficient manner. The reported results demonstrate that the system achieves high specificity and sensitivity in detecting sepsis episodes, accurately identifying their onset and duration. This system has multiple applications, including potentially supporting clinical decision-making in the PICU, labeling large datasets for developing predictive models, and facilitating automated surveillance of sepsis and quality management in pediatric critical care.

## Data Availability

Data access can be requested upon purely scientific purpose [31].
All DTA analyses were conducted in R version 4.4.0 (2024-04-24). The corresponding scripts can be accessed via https://zivgitlab.uni-muenster.de/boehnkej/elise-dta-sirs-models-sepsis-models

https://zivgitlab.uni-muenster.de/boehnkej/elise-dta-sirs-models-sepsis-models

## LIST OF ABBREVIATIONS

ICD: International Classification of Diseases
CDSS: Clinical Decision Support System
CMV: Cytomegalovirus
CRP: C-reactive protein
DNA: Deoxyribonucleic acid
DTA: Diagnostic Test Accuracy
EBV: Epstein–Barr virus
FN: False Negative
FP: False Positive
IL6: Interleukin 6
IPSCC: International Pediatric Sepsis Consensus Conference
MHH: Hannover Medical School
OPS: Operationen-und Prozedurenschlüssel (Operation and Procedure Classification System),
PCR: Polymerase chain reaction
PICU: Pediatric Intensive Care Unit
SIRS: Systemic Inflammatory Response Syndrome

## DECLARATIONS

### Ethics approval

All study participants, their parents, or legal guardians gave written informed consent. The study has been approved by the Ethics Committee of Hannover Medical School (No. 7804_BO_S_2018 and No. 9819_BO_S_2021). The study was performed in compliance with the World Medical Association Declaration of Helsinki on Ethical Principles for Medical Research Involving Human Subjects and was reviewed by the Ethics Committee of Hannover Medical School.

### Consent for publication

Not applicable

### Availability of data and materials

Data access can be requested upon purely scientific purpose [31].

All DTA analyses were conducted in R version 4.4.0 (2024-04-24). The corresponding scripts can be accessed via https://zivgitlab.uni-muenster.de/boehnkej/elise-dta-sirs-models-sepsis-models.

### Authors’ contributions

All authors were involved in the acquisition of data, knowledge, or rules and in the revision of the manuscript. They approved the final version.

M.S. was responsible for organizing the drafting process and manuscript writing, developing the CDSS rules, development of the open demonstrator, and outlining the manuscript.

H.R. was responsible for organizing the drafting process and manuscript writing, evaluating the patients to define the reference standard decisions, and outlining the manuscript.

J.B. was responsible for conducting the statistical analysis and Proof-of-Concept DTA evaluation.

N.R. was responsible for conducting the statistical analysis and Proof-of-Concept DTA evaluation.

L.B. was responsible for the development of the CDSS rules.

A.K. was responsible for the statistical analysis and Proof-of-Concept DTA evaluation.

M.A. was responsible for the development of the open demonstrator.

M.M. supervised the work, critically revised the manuscript, and gave further methodological advice.

P.B. supervised the work, critically revised the manuscript, and gave further methodological advice.

A.W. designed the study, provided expertise and data, was responsible for the development of the CDSS rules and overall CDSS architecture, and co-drafted the manuscript.

T.J. designed the study, provided clinical expertise and data, evaluated the patients to define the reference standard for final decisions, and co-drafted the manuscript.

The authors used generative AI and AI-assisted technologies in the writing process to improve readability and language of the work. They applied the technology with human oversight and control. All work was reviewed and edited carefully.

## Acknowledgements

We would like to thank the ELISE Study Group for its input. Moreover, the assistance provided by the MHH Information Technology is greatly appreciated. The authors acknowledge the support of the Ministry of Science and Culture of Lower Saxony through funds from the program zukunft.niedersachsen of the Volkswagen Foundation for the ‘CAIMed – Lower Saxony Center for Artificial Intelligence and Causal Methods in Medicine’ project (grant no. ZN4257).

## Footnotes

2 https://medisite.net/

3 www.github.com/ehrbase

4 https://gwdg.de/en/services/server-services/gwdg-cloud-server

