## Supplements for "An Open Demonstrator for an Interoperable Clinical Decision Support System for the Detection of Systemic Inflammation and Sepsis in Pediatric Intensive Care"

### Appendix A: DTAs of the sepsis detection models

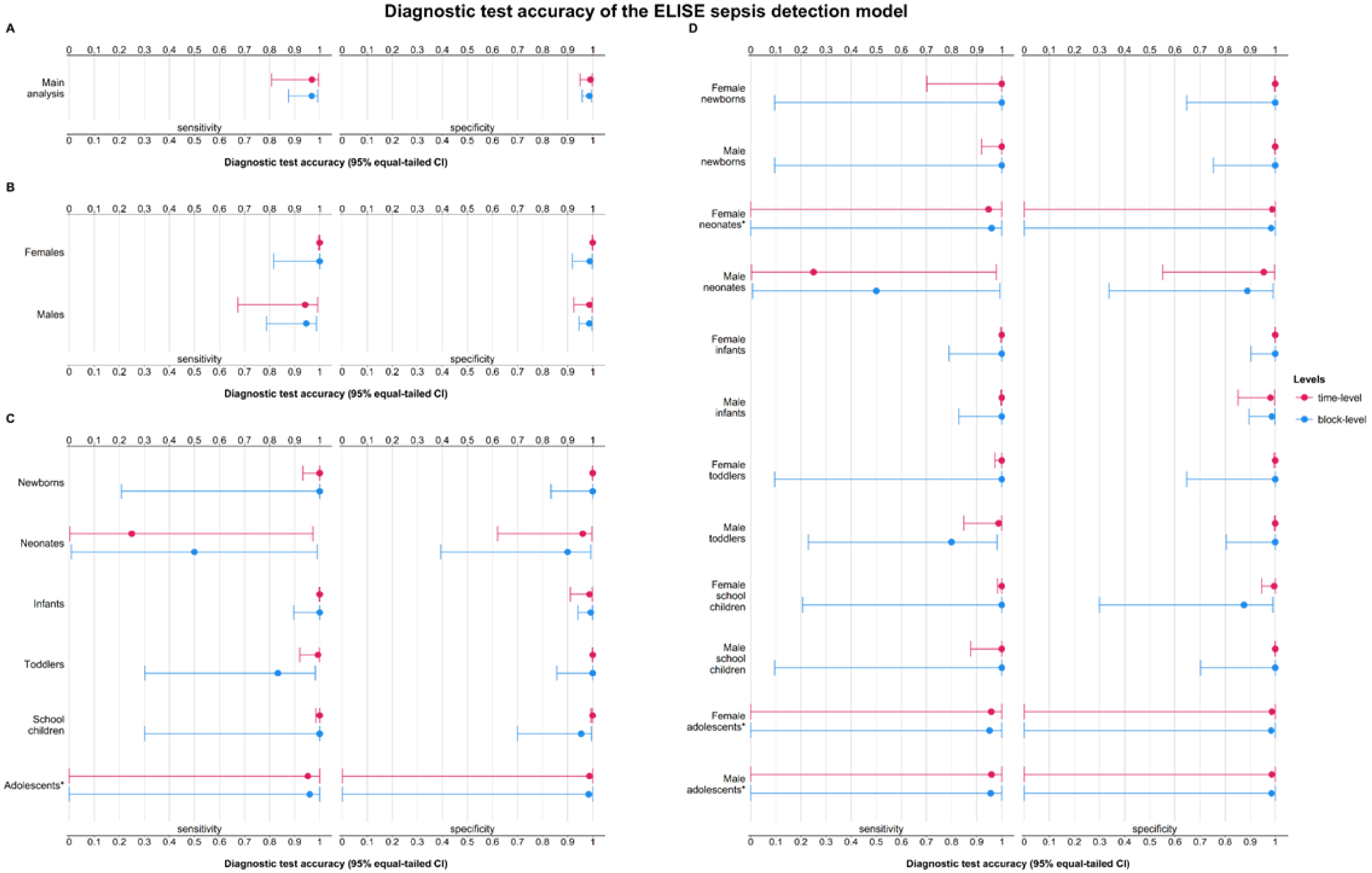

**Figure B.1:** This figure presents the diagnostic test accuracy (DTA) of the sepsis detection model (index test) in comparison to the clinician's diagnostic decision. The evaluation of the primary analysis (= the overall performance of the index test) is presented in Plot A. To facilitate deeper exploration, Plots B to D devel into performance differences via stratified analyses. Specifically, Plot B presents the index tests DTA per time point stratified by sex, Plot C stratified by age groups, and Plot D stratified by sex per age group. Whenever there were no false positive or false negative results (i.e. the point estimate was 100%), we used Wilson's method to estimate a lower bound of DTA ignoring the data clustering (i.e. which DTA could have been achieved if one false result had had occurred).

* Sensitivity and specificity were estimated via a beta distribution since there were either no true positive or no true negative results.

| Diagnosis | Population | Estimation level | DTA metrics | | Contingency table | | | |
|---|---|---|---|---|---|---|---|---|
| | | | Sensitivity (95% CI) | Specificity (95% CI) | True Positive* | False Positive* | False Negative* | True Negative* |
| Sepsis (proven/suspected) *Time unit: per hour* | All patients | Time-level | 96.9% (80.9-99.6) | 99.1% (95.1-99.8%) | 3,450 | 386 | 111 | 43,594 |
| | | Block-level | 96.8% (87.5-99.3%) | 98.6% (95.8-99.6%) | 61 | 3 | 2 | 219 |
| Sepsis (proven/suspected) *Time unit: per hour* | Females | Time-level | 100% (min. 99.7%)† | 100% (99.8-100%) | 1,662 | 3 | 0 | 13,954 |
| | | Block-level | 100% (min. 81.7%)† | 98.8% (91.8-99.8%) | 26 | 1 | 0 | 85 |
| | Males | Time-level | 94.2% (67.3-99.2%) | 98.7% (92.4-99.8%) | 1,788 | 383 | 111 | 29,640 |
| | | Block-level | 94.6% (78.7-98.8%) | 98.6% (94.6-99.7%) | 35 | 2 | 2 | 144 |
| Sepsis (proven/suspected) *Time unit: per hour* | Newborns | Time-level | 100% (min. 93.3%)† | 100% (min. 99.9%)† | 79 | 0 | 0 | 6,052 |
| | | Block-level | 100% (min. 20.8%)† | 100% (min. 83.3%)† | 2 | 0 | 0 | 29 |
| | Neonates | Time-level | 25.0% (0.3-94.4%) | 96.1% (62.1-99.7%) | 36 | 47 | 108 | 1,148 |
| | | Block-level | 50.0% (0.9-99.1%) | 90.0% (39.4-99.2%) | 1 | 1 | 1 | 9 |
| | Infants | Time-level | 100% (min. 99.8%)† | 98.7% (91.1-99.8%) | 2,521 | 336 | 0 | 26,493 |
| | | Block-level | 100% (89.7-100%)† | 99.2% (94.2-99.9%) | 50 | 1 | | 125 |
| | Toddlers | Time-level | 99.3% (92.1-99.9%) | 100% (min. 99.9%)† | 445 | 0 | 3 | 3,997 |
| | | Block-level | 83.3% (30.2-98.3%) | 100% (min. 85.8%)† | 5 | 0 | 1 | 35 |
| | School children | Time-level | 100% (min. 98.5%)† | 99.9% (99.3-100%) | 369 | 3 | 0 | 4,719 |
| | | Block-level | 100% (min. 30.1%)† | 95.5% (70.1-99.5%) | 3 | 1 | 0 | 21 |
| | Adolescents‡ | Time-level | 95.2% | 98.7% | 0 | 0 | 0 | 1,185 |

| Diagnosis | Population | Estimation level | DTA metrics | | Contingency table | | | |
|---|---|---|---|---|---|---|---|---|
| | | | Sensitivity (95% CI) | Specificity (95% CI) | True Positive* | False Positive* | False Negative* | True Negative* |
| | | | (min. 0%)† | (min. 0%)† | | | | |
| | | Block-level | 96.0% (min. 0%)† | 98.4% (min. 0%)† | 0 | 0 | 0 | 10 |
| Sepsis (proven/suspected) *Time unit: per hour* | Female newborns | Time-level | 100% (min. 70.2%)† | 100% (min. 99.8%)† | 14 | 0 | 0 | 2,463 |
| | | Block-level | 100% (min. 9.5%)† | 100% (min. 64.7%)† | 1 | 0 | 0 | 11 |
| | Male newborns | Time-level | 100% (min. 91.9%)† | 100% (min. 99.8%)† | 65 | 0 | 0 | 3,589 |
| | | Block-level | 100% (min. 9.5%)† | 100% (min. 75.4%)† | 1 | 0 | 0 | 18 |
| | Female neonates‡ | Time-level | 94.8% (min. 0%)† | 98.8% (min. 0%)† | 0 | 0 | 0 | 184 |
| | | Block-level | 96.0% (min. 0%)† | 98.3% (min. 0%)† | 0 | 0 | 0 | 1 |
| | Male neonates | Time-level | 25.0% (0.2-97.8%) | 95.4% (55.1-99.7%) | 36 | 47 | 108 | 964 |
| | | Block-level | 50.0% (0.7-99.3%) | 88.9% (33.8-99.2%) | 1 | 1 | 1 | 8 |
| | Female infants | Time-level | 100% (min. 99.5%)† | 100% (min. 99.9%)† | 1,112 | 0 | 0 | 9,430 |
| | | Block-level | 100% (min. 79.0%)† | 100% (min. 90.4%)† | 22 | 0 | 0 | 54 |
| | Male infants | Time-level | 100% (min. 99.6%)† | 98.1% (85.2-99.8%) | 1,409 | 336 | 0 | 17,063 |
| | | Block-level | 100% (min. 82.8%)† | 98.6% (89.6-99.8%) | 28 | 1 | 0 | 71 |
| | Female toddlers | Time-level | 100% (min. 97.3%)† | 100% (min. 99.5%)† | 208 | 0 | 0 | 1,084 |
| | | Block-level | 100% (min. 9.5%)† | 100% (min. 64.6%)† | 1 | 0 | 0 | 11 |
| | Male toddlers | Time-level | 98.8% (84.9-99.9%) | 100% (min. 99.8%)† | 237 | 0 | 3 | 2,913 |

| Diagnosis | Population | Estimation level | DTA metrics | | Contingency table | | | |
|---|---|---|---|---|---|---|---|---|
| | | | Sensitivity (95% CI) | Specificity (95% CI) | True Positive* | False Positive* | False Negative* | True Negative* |
| | | Block-level | 80.0% (22.9-98.2%) | 100% (min. 80.5%)† | 4 | 0 | 1 | 24 |
| | Female school children | Time-level | 100% (min. 98.3%)† | 99.6% (94.6-100%) | 328 | 3 | 0 | 743 |
| | | Block-level | 100% (min. 20.8%)† | 87.5% (29.9-99.1%) | 2 | 1 | 0 | 7 |
| | Male school children | Time-level | 100% (min. 87.7%)† | 100% (min. 99.9%)† | 41 | 0 | 0 | 3,976 |
| | | Block-level | 100% (min. 9.5%)† | 100% (min. 70.2%)† | 1 | 0 | 0 | 14 |
| | Female adolescents‡ | Time-level | 95.8% (min. 0%)† | 98.6% (min. 0%)† | 0 | 0 | 0 | 50 |
| | | Block-level | 95.2% (min. 0%)† | 98.3% (min. 0%)† | 0 | 0 | 0 | 1 |
| | Male adolescents‡ | Time-level | 95.9% (min. 0%)† | 98.6% (min. 0%)† | 0 | 0 | 0 | 1,135 |
| | | Block-level | 95.6% (min. 0%)† | 98.5% (min. 0%)† | 0 | 0 | 0 | 9 |

**Table A.1:** This table presents the diagnostic test accuracy (DTA) of the sepsis detection model (index test) as compared to the clinician's diagnostic decision, which defines the ground truth.DTA = diagnostic test accuracy. **Abbreviations**: CI = confidence interval

* all time points in the dataset with the corresponding label (i.e., multiple time points per patient)

† we used Wilson's method to estimate a lower bound of DTA ignoring the data clustering (i.e. which DTA could have been achieved if one false result had had occurred)

‡ sensitivity and specificity were estimated via a beta distribution since either no true positive or true negative classification labels are present. As parameters for the beta distribution, we took 100 random samples to estimate the median sensitivity and specificity. This approach is proposed by Lash, Fox and Fink [34].

3

**Appendix B: Adapted Clinical Decision Support System for SIRS Detection**

As part of the adaptation of the knowledge representation and rule development we adapted or introduced a total of eleven rules, summarized in Table A1. From a close cooperation with experienced pediatricians and the observation of the clinical routine, we derived various rules that relate to all four criteria of the SIRS diagnostic. We have four rule changes (T1 to T4) for the temperature criterion. The T1 rule adjusts the hypothermia threshold for patients younger than one year to 35.7°C, while patients older than one year cannot have hypothermia by definition. In addition, abnormal temperature values during temperature regulation (T2), within four hours after surgery or procedure (T3) or within the first six hours after admission (T4) are not considered relevant for triggering SIRS. These rules stem from the fact that patients can become hypothermic or have an elevated temperature due to stress after being transferred or transported through the hospital. However, this temperature fluctuation is not triggered by SIRS and is therefore not relevant for the diagnosis. For the criterion of respiratory rate, we adapted one rule. Abnormal respiratory rates during mechanical ventilation are only considered as a SIRS trigger when the respiratory rate median is 1.5 times higher than the set respiratory rate during mechanical ventilation. Regarding leukocyte counts, we adjusted three rules. First, analogues to leukocyte count, a high number of immature granulocytes are considered a SIRS trigger (L1). To avoid false positive leukopenia, a low leukocyte count is only considered a SIRS trigger in the absence of documented diagnoses of agranulocytosis. Likewise, leukocytosis is only used as a SIRS trigger if no non-follicular lymphomata, types of non-Hodgkin lymphomata or certain leukemia is documented. For the last criterion of heart rate, we made three further adjustments. Bradycardia is only considered a SIRS trigger if no beta-blockers were previously administered (HR1, HR2). As a final adjustment, abnormal heart rates are not considered when a pacemaker is active (HR3).

| | Rule Name | Value | Adaption | Start of criterion | End of criterion |
|---|---|---|---|---|---|
| Temperature | T1 | Median of temperature values of the last hour | The median value is only considered as too low when below 35.7°C and only when the patient is younger than 365 days. | One hour before the timestamp of temperature measurement | One and a half hours after the timestamp of measurement |
| | T2 | Median of temperature values of the last hour | When an active temperature regulation is applied, too low or too high temperature medians are not considered as SIRS criterion. | | |
| | T3 | Median of temperature values of the last hour | During 4 hours after a procedure a too low temperature median is not considered as a SIRS criterion. | | |
| | T4 | Median of temperature values of the last hour | During 6 hours after an admission to the ward a too low temperature median is not considered as a SIRS criterion. | | |
| Respiratory Rate | RR1 | Median of respiratory rate values of the last hour | During an applied mechanical ventilation the value is only considered as too high when the median value is 1.5 times higher than the setting parameter of the mechanical ventilation | One hour before the timestamp of respiratory rate value | Timestamp of respiratory rate value |
| Leukocyte Count | L1 | Laboratory test results involving immature granulocytes | Analogous to leukocyte counts immature granulocytes are considered as a SIRS criterion when higher than 10% | Timestamp of the collection of the specimen. | 24 hours after the timestamp of the collection of the specimen |

| | | | |
|---|---|---|---|
| | L2 | Laboratory test results involving leukocyte count | A too low leukocyte count is only valued when none of the following ICD-10 diagnoses have been documented: D70.0, D70.1, D70.3, D70.5, D70.6, D70.7, D76.1 |
| | L3 | Laboratory test results involving leukocyte count | A high low leukocyte count is only valued when none of the following ICD-10 diagnoses have been documented: C83.0, C83.1, C83.5, C83.7, C85.1, C85.2, C85.7, C85.9, C91.0, C91.1, C91.3, C91.4, C91.5, C91.6, C91.7, C91.8, C91.9, C92.0, C92.1, C92.2, C92.3, C92.4, C92.5, C92.6, C92.7, C92.8, C92.9, C93.0, C93.1, C93.3, C93.7, C93.9, C95.0, C95.1, C95.7, C95.8, C95.9 |
| Heart Rate | HR1 | Median of heart rate values of the last hour | The median value is only considered as too low when in the previous 24 hours no beta-blocker (Clonidin, Propranolol, Metoprolol, Bisoprolol, Esmolol or Landiolol) has been given. |
| | HR2 | Median of heart rate values of the last hour | The median value is only considered as too low when in the previous 72 hours no beta-blocker (Amidoran) has been given. |
| | HR3 | Median of heart rate values of the last hour | When an active pacer is applied a too high heart rate median is not considered as a SIRS criterion. |

**Table B1.** A summary of the rules that are currently in place within the clinical decision support system, which relate to the diagnosis of Systemic inflammatory response syndrome. **Abbreviations:** ICD, International Classification of Diseases; SIRS, Systemic inflammatory response syndrome.

### Appendix C: Diagnostic test accuracies of the SIRS detection models

**Objective:**

This evaluation aims to assess the diagnostic test accuracy (DTA) of two Systemic Inflammatory Response Syndrome (SIRS) detection models: the published CADDIE SIRS model developed by Wulff et al. [22] and the ELISE SIRS model. The ELISE SIRS model is an updated version of the CADDIE SIRS model that incorporates new knowledge in developing the decision rules.

**Methodology:**

For each index test (i.e. the SIRS detection models), we merged patient data (age, sex, and observation periods, i.e. length of stay at PICU) with start and end times of episodes according to reference standard (clinician's diagnoses blinded to index test results) and respective index test. Per patient, any episodes less than 36 hours apart were merged into one continuing episode. There were neither missing nor indetermined test results.

We estimated DTA metrics—sensitivity, specificity, and their 95% Wald confidence intervals (where possible)—with the method by Böhnke et al. [26] Using hour as time unit, we estimated the DTA at both the time-level (measuring the model's precision in identifying correct time points) and block-level (evaluating performance over clinically relevant time periods). For the block-level, the time point labels per block were summarized into one single label; false positive and false negative labels overrule true positive and true negative labels. This labelling penalized any differences between the diagnostic tests, e.g. if the index test detects SIRS just one hour before the reference standard, which would still be correct in clinical practice. We applied two modifying rules to estimate a clinically relevant DTA: 1) We applied a clinician-based tolerance margin rule so that if the index test starts/ends within the tolerance margin of the reference standard, the index test's diagnostic status of the specific time points is changed in accordance with the reference standard's diagnostic status (i.e., no "punishment" if index test starts/ends too early/late); 2) A %-correctness rule was applied according to which the index test's diagnostic status per patient is corrected in accordance with the reference standard's diagnostic status if at minimum Pdiseased% of single time points per a diseased block and at minimum Pdisease-free% of single time points per a disease-free block are correctly classified. For our analysis, we used a tolerance margin of ±4 hours around the reference standard's disease episode start and end; and a 75%-correction rule for diseased and disease-free blocks.

Whenever there were no false positive or false negative results so that we could not estimate a 95% Wald confidence interval, we used Wilson's method [32] to estimate a lower bound of DTA ignoring the data clustering (i.e. which DTA could have been achieved if one false result had occurred). We conducted exploratory stratified analyses for sex, age groups, and all combinations of sex by age groups, using the same methods as the main analyses.

Given that both detection models detect the same target condition, we also performed a comparison of the two models using McNemar's test.

**Results***:*

Irrespective of the estimation level, the ELISE SIRS model exhibited lower sensitivities, yet higher specificities compared to the CADDIE SIRS model (Figure C.1 and Table C.1): On the time level, the ELISE SIRS model detected the presence of SIRS with a sensitivity of 87.9% (CADDIE SIRS model: 92.4%) and the absence of SIRS with a specificity of 97.1% (CADDIE SIRS model: 92.3%). On the block-level after applying the modifying rules, the ELISE SIRS model detected SIRS events with a sensitivity of 81.9% (CADDIE SIRS model:

84.2%) and disease-free periods with a specificity of 96.4% (CADDIE SIRS model: 88.6%).

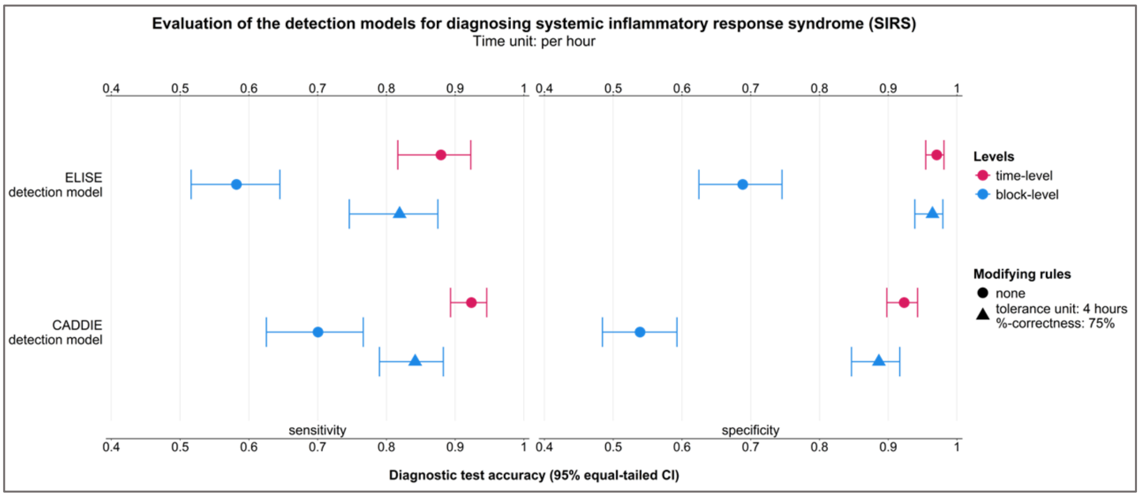

**Figure C.2:** Diagnostic test accuracy estimations of the SIRS detection models (index tests) per estimation level using an hourly time unit.

| | Estimation level | DTA metrics | | Contingency table | | | |
|---|---|---|---|---|---|---|---|
| | | Sensitivity (95% CI) | Specificity (95% CI) | True Positive* | False Positive* | False Negative* | True Negative* |
| SIRS ELISE model *Time unit: per hour* | Time-level | 87.9% (81.7-92.3%) | 97.1% (95.5-98.1%) | 6,415 | 1,186 | 880 | 39,060 |
| | Block-level† | 58.2% (51.6-64.5%) | 68.8% (62.5-74.6%) | 103 | 96 | 74 | 212 |
| | Block-level‡ | 81.9% (74.6-87.5%) | 96.4% (93.9-97.9%) | 145 | 11 | 32 | 297 |

| | Estimation level | DTA metrics | | Contingency table | | | |
|---|---|---|---|---|---|---|---|
| | | Sensitivity (95% CI) | Specificity (95% CI) | True Positive* | False Positive* | False Negative* | True Negative* |
| SIRS CADDIE model *Time unit: per hour* | Time-level | 92.4% (89.3-94.6%) | 92.3% (89.8-94.3%) | 6,738 | 3,089 | 557 | 37,157 |
| | Block-level† | 70.1% (62.5-76.6%) | 53.9% (48.4-59.3%) | 124 | 142 | 53 | 166 |
| | Block-level‡ | 84.2% (79.0-88.3%) | 88.6% (84.7-91.7%) | 149 | 35 | 28 | 273 |

**Table C.2:** Diagnostic test accuracy estimations of the SIRS detection models (index tests) per estimation level using an hourly time unit. **Abbreviations**: DTA = diagnostic test accuracy; CI = confidence interval; SIRS = systemic inflammatory response syndrome

* all time points in the dataset with the corresponding label (i.e., multiple time points per patient)

† estimates without modifying rules

‡ estimates using a tolerance window of 4 hours around the start and end of the reference standard episode and 75%-correctness rule per block

The stratified DTA evaluations showed that the models performed less well in certain clusters (i.e., newborns, neonates, and adolescents regardless of sex; Figure C.2 and C.3).

The comparison of the two models (Table C.2) confirmed that both SIRS detection models are statistically significantly different (p-value <0.001 in both McNemar's tests).

| **Clinician's diagnosis: SIRS present** | **CADDIE model** | | **Total** |
|---|---|---|---|
| | SIRS detected | No SIRS detected | |

| | | | | |
|---|---|---|---|---|
| **ELISE model** | SIRS detected | 6,137 | 278 | 6,415 |
| | No SIRS detected | 601 | 279 | 880 |
| | **Total** | 6,738 | 557 | 7,295 |
| **Clinician's diagnosis: SIRS absent** | | **CADDIE model** | | **Total** |
| | | SIRS detected | No SIRS detected | |
| **ELISE model** | SIRS detected | 842 | 344 | 1,186 |
| | No SIRS detected | 2,247 | 36,813 | 39,060 |
| | **Total** | 3,089 | 37,157 | 40,246 |

**Table C.2**: 2×2 contingency tables to perform the McNemar's tests.

**Conclusion:**

Both SIRS detection models can support the diagnostic decision-making process in clinical practice. It is up to the clinicians to decide which to use depending on the trade-off between sensitivity and specificity.

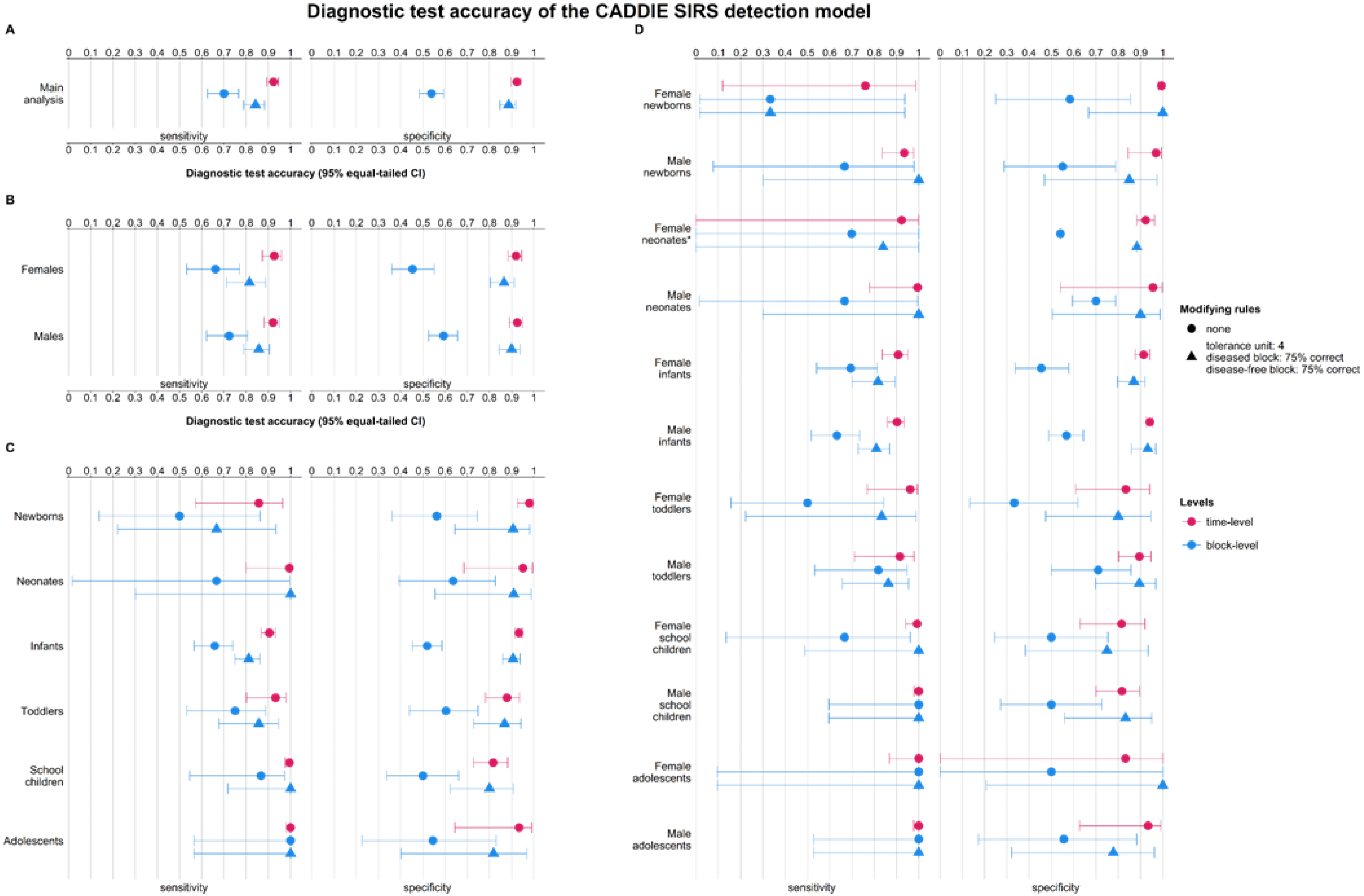

**Figure C.3:** This figure presents the diagnostic test accuracy (DTA) of the CADDIE SIRS detection model (index test) in comparison to the clinician's diagnostic decision which defines the ground truth. The evaluation of the primary analysis (= the overall performance of the index test) is presented in Plot A. To facilitate deeper exploration, Plots B to D devel into performance differences via stratified analyses. Specifically, Plot B presents the index tests DTA per time point stratified by sex, Plot C stratified by age groups, and Plot D stratified by sex per age group. Whenever there were no false positive or false negative results (i.e. the point estimate was 100%), we used Wilson's method to estimate a lower bound of DTA ignoring the data clustering (i.e. which DTA could have been achieved if one false result had had occurred).

* Sensitivity and specificity were estimated via a beta distribution since there were either no true positive or no true negative results.

| Diagnosis | Population | Estimation level | DTA metrics: Sensitivity (95% CI) | DTA metrics: Specificity (95% CI) | Contingency table: True Positive* | Contingency table: False Positive* | Contingency table: False Negative* | Contingency table: True Negative* |
|---|---|---|---|---|---|---|---|---|
| CAD DIE SIRS model *Time unit: per hour* | Females | Time-level | 92.7% (87.3-95.9%) | 92.0% (88.5-94.5%) | 2,863 | 1,006 | 226 | 11,524 |
| | | Block-level† | 66.2% (53.2-77.1%) | 45.4% (36.0-55.1%) | 43 | 65 | 22 | 54 |
| | | Block-level‡ | 81.5% (71.2-88.7%) | 86.6% (80.4-91.0%) | 53 | 16 | 12 | 103 |
| | Males | Time-level | 92.1% (88.2-94.8%) | 92.5% (89.0-94.9%) | 3,875 | 2,083 | 331 | 25,633 |
| | | Block-level† | 72.3% (62.2-80.6%) | 59.3% (52.6-65.7%) | 81 | 81 | 31 | 118 |
| | | Block-level‡ | 85.7% (79.1-90.5%) | 89.9% (84.2-93.7%) | 96 | 20 | 16 | 179 |
| CAD DIE SIRS model | Newborns | Time-level | 85.7% (57.2-96.4%) | 98.0% (92.7-99.5%) | 144 | 121 | 24 | 5,842 |

| Diagnosis | Population | Estimation level | DTA metrics: Sensitivity (95% CI) | DTA metrics: Specificity (95% CI) | Contingency table: True Positive* | Contingency table: False Positive* | Contingency table: False Negative* | Contingency table: True Negative* |
|---|---|---|---|---|---|---|---|---|
| *Time unit: per hour* | | Block-level† | 50.0% (13.7-86.3%) | 56.2% (36.1-74.5%) | 3 | 14 | 3 | 18 |
| | | Block-level‡ | 66.7% (22.2-93.4%) | 90.6% (64.5-98.1%) | 4 | 3 | 2 | 29 |
| | Neonates | Time-level | 99.5% (80.0-100%) | 90.5% (68.6-99.4%) | 190 | 57 | 1 | 1,091 |
| | | Block-level† | 66.7% (1.7-99.6%) | 63.6% (39.2-82.6%) | 2 | 4 | 1 | 7 |
| | | Block-level‡ | 100% (min. 30.1%)§ | 90.9% (55.5-98.8%) | 3 | 1 | 0 | 10 |
| | Infants | Time-level | 90.5% (86.9-93.1%) | 93.3% (91.5-94.7%) | 4,347 | 1,654 | 458 | 22,891 |
| | | Block-level† | 65.8% (56.6-74.0%) | 51.9% (45.2-58.6%) | 77 | 87 | 40 | 94 |

| Diagnosis | Population | Estimation level | DTA metrics | | Contingency table | | | |
|---|---|---|---|---|---|---|---|---|
| | | | Sensitivity (95% CI) | Specificity (95% CI) | True Positive* | False Positive* | False Negative* | True Negative* |
| | | Block-level‡ | 81.2% (74.8-86.2%) | 90.6% (85.9-93.8%) | 95 | 17 | 22 | 164 |
| | Toddlers | Time-level | 93.2% (80.2-97.9%) | 87.9% (78.4-93.6%) | 966 | 412 | 70 | 2,997 |
| | | Block-level† | 75.0% (53.3-88.7%) | 60.4% (43.9-74.8%) | 21 | 21 | 7 | 32 |
| | | Block-level‡ | 85.7% (67.8-94.5%) | 86.8% (72.7-94.2%) | 24 | 7 | 4 | 46 |
| | School children | Time-level | 99.5% (97.6-99.9%) | 81.7% (72.7-88.2%) | 802 | 785 | 4 | 3,500 |
| | | Block-level† | 86.7% (54.5-97.2%) | 50.0% (33.9-66.1%) | 13 | 15 | 2 | 15 |
| | | Block-level‡ | 100% (min. 71.7%)§ | 80.0% (62.2-90.7%) | 15 | 6 | 0 | 24 |

| Diagnosis | Population | Estimation level | DTA metrics: Sensitivity (95% CI) | DTA metrics: Specificity (95% CI) | Contingency table: True Positive* | Contingency table: False Positive* | Contingency table: False Negative* | Contingency table: True Negative* |
|---|---|---|---|---|---|---|---|---|
| | Adolescents | Time-level | 100% (min. 98.1%)§ | 93.3% (64.5-99.1%) | 289 | 60 | 0 | 836 |
| | | Block-level† | 100% (min. 56.5%)§ | 54.5% (22.7-83.0%) | 8 | 5 | 0 | 6 |
| | | Block-level‡ | 100% (min. 56.5%)§ | 81.8% (40.1-96.8%) | 8 | 2 | 0 | 9 |
| CADDIE SIRS model *Time unit: per hour* | Female newborns | Time-level | 76.0% (11.9-98.7%) | 99.4% (98.0-99.8%) | 57 | 14 | 18 | 2,388 |
| | | Block-level† | 33.3% (1.7-93.7%) | 58.3% (24.9-85.5%) | 1 | 5 | 2 | 7 |
| | | Block-level‡ | 33.3% (1.7-93.7%) | 100% (min. 66.7%)§ | 1 | 0 | 2 | 12 |
| | Male newborns | Time-level | 93.5% (83.5-97.7%) | 97.0% (84.4-99.5%) | 87 | 107 | 6 | 3,454 |

| Diagnosis | Population | Estimation level | DTA metrics | | Contingency table | | | |
|---|---|---|---|---|---|---|---|---|
| | | | Sensitivity (95% CI) | Specificity (95% CI) | True Positive* | False Positive* | False Negative* | True Negative* |
| | | Block-level† | 66.7% (7.7-98.0%) | 55.0% (28.8-78.7%) | 2 | 9 | 1 | 11 |
| | | Block-level‡ | 100% (min. 30.1%)§ | 85.0% (46.9-97.3%) | 3 | 3 | 0 | 17 |
| | Female neonates¶ | Time-level | 92.3% (min. 0%)§ | 92.3% (min. 0%)§ | 0 | 15 | 0 | 169 |
| | | Block-level† | 70.2% (min. 0%)§ | 54.0% (min. 0%)§ | 0 | 1 | 0 | 0 |
| | | Block-level‡ | 84.0% (min. 0%)§ | 88.4% (min. 0%)§ | 0 | 0 | 0 | 1 |
| | Male neonates | Time-level | 99.5% (77.8-100%) | 95.6% (54.1-99.8%) | 190 | 42 | 1 | 922 |
| | | Block-level† | 66.7% (1.5-99.6%) | 70.0% (59.4-78.8%) | 2 | 3 | 1 | 7 |
| | | Block-level‡ | 100% (min. 30.1%)§ | 90.0% | 3 | 1 | 0 | 9 |

| Diagnosis | Population | Estimation level | DTA metrics: Sensitivity (95% CI) | DTA metrics: Specificity (95% CI) | Contingency table: True Positive* | Contingency table: False Positive* | Contingency table: False Negative* | Contingency table: True Negative* |
|---|---|---|---|---|---|---|---|---|
| | | | | (50.6-98.8%) | | | | |
| | Female infants | Time-level | 90.8% (83.6-95.0%) | 91.4% (87.5-94.2%) | 1,859 | 727 | 189 | 7,767 |
| | | Block-level† | 69.4% (54.1-81.3%) | 45.5% (33.7-57.7%) | 34 | 42 | 15 | 35 |
| | | Block-level‡ | 81.6% (69.9-89.5%) | 87.0% (79.7-92.0%) | 40 | 10 | 9 | 67 |
| | Male infants | Time-level | 90.2% (86.0-93.3%) | 94.2% (92.6-95.5%) | 2,488 | 927 | 269 | 15,124 |
| | | Block-level† | 63.2% (51.6-73.5%) | 56.7% (48.9-64.3%) | 43 | 45 | 25 | 59 |
| | | Block-level‡ | 80.9% (72.8-87.0%) | 93.3% (86.0-96.9%) | 55 | 7 | 13 | 97 |
| | Female | Time-level | 96.2% | 83.5% | 375 | 149 | 15 | 753 |

| Diagnosis | Population | Estimation level | DTA metrics: Sensitivity (95% CI) | DTA metrics: Specificity (95% CI) | Contingency table: True Positive* | Contingency table: False Positive* | Contingency table: False Negative* | Contingency table: True Negative* |
|---|---|---|---|---|---|---|---|---|
| | toddlers | | (76.7-99.5%) | (61.0-94.2%) | | | | |
| | | Block-level† | 50.0% (15.6-84.4%) | 33.3% (13.4-61.8%) | 3 | 10 | 3 | 5 |
| | | Block-level‡ | 83.3% (22.2-98.9%) | 80.0% (47.4-94.7%) | 5 | 3 | 1 | 12 |
| | Male toddlers | Time-level | 91.5% (71.0-97.9%) | 89.5% (80.2-94.7%) | 591 | 263 | 55 | 2,244 |
| | | Block-level† | 81.8% (53.3-94.7%) | 71.1% (50.1-85.7%) | 18 | 11 | 4 | 27 |
| | | Block-level‡ | 86.4% (65.7-95.4%) | 89.5% (69.9-96.9%) | 19 | 4 | 3 | 34 |
| | Female school children | Time-level | 99.3% (94.2-99.9%) | 81.5% (62.8-92.0%) | 534 | 99 | 4 | 437 |
| | | Block-level† | 66.7% | 50.0% | 4 | 6 | 2 | 6 |

| Diagnosis | Population | Estimation level | DTA metrics | | Contingency table | | | |
|---|---|---|---|---|---|---|---|---|
| | | | Sensitivity (95% CI) | Specificity (95% CI) | True Positive* | False Positive* | False Negative* | True Negative* |
| | | | (13.4-96.3%) | (24.5-75.5%) | | | | |
| | | Block-level‡ | 100% (min. 48.7%)§ | 75.0% (38.3-93.6%) | 6 | 3 | 0 | 9 |
| | Male school children | Time-level | 100% (min. 97.9%)§ | 81.7% (70.0-89.5%) | 268 | 686 | 0 | 3,063 |
| | | Block-level† | 100% (min. 59.6%)§ | 50.0% (27.2-72.8%) | 9 | 9 | 0 | 9 |
| | | Block-level‡ | 100% (min. 59.6%)§ | 83.3% (55.9-95.2%) | 9 | 3 | 0 | 15 |
| | Female adolescents | Time-level | 100% (min. 86.8%)§ | 83.3% (min. 0%)§ | 38 | 2 | 0 | 10 |
| | | Block-level† | 100% (min. 9.5%)§ | 50.0% (min. 0%)§ | 1 | 1 | 0 | 1 |
| | | Block-level‡ | 100% (min. 9.5%)§ | 100% (min. 20.8%)§ | 1 | 0 | 0 | 2 |

| Diagnosis | Population | Estimation level | DTA metrics | | Contingency table | | | |
|---|---|---|---|---|---|---|---|---|
| | | | Sensitivity (95% CI) | Specificity (95% CI) | True Positive* | False Positive* | False Negative* | True Negative* |
| | Male adolescents | Time-level | 100% (min. 97.8 %)[§] | 93.4 % (62.7-99.2 %) | 251 | 58 | 0 | 826 |
| | | Block-level[†] | 100% (min. 52.9 %)[§] | 55.6 % (17.2-88.3 %) | 7 | 4 | 0 | 5 |
| | | Block-level[‡] | 100% (min. 52.9 %)[§] | 77.8 % (32.2-96.3 %) | 7 | 2 | 0 | 7 |

**Table C.3**: Exploratory diagnostic test accuracy estimations of the CADDIE SIRS detection model (index test) developed by Wulff et al. 1 per estimation level using an hourly time unit. **Abbreviations**: CI = confidence interval; SIRS = systemic inflammatory response syndrome

* all time points in the dataset with the corresponding label (i.e., multiple time points per patient)

† naïve estimates without adjustments

‡ adjusted estimates using a $t_{tolerance\ window}$ of 4 hours around the start and end of the reference standard episode and 75%-correctness rule per block

§ we used Wilson's method to estimate a lower bound of DTA ignoring the data clustering (i.e. which DTA could have been achieved if one false result had had occurred)

¶ sensitivity and specificity were estimated via a beta distribution since either no true positive or true negative classification labels are present. As parameters for the beta distribution, we took 100 random samples to estimate the median sensitivity and specificity. This approach is proposed by Lash, Fox and Fink [34]
